# Metabolic indicators associated with non-communicable diseases deteriorated in COVID-19 outbreak: evidence from a two-center, retrospective study

**DOI:** 10.1101/2020.07.02.20144857

**Authors:** Ting Xue, Lizhen Xu, Yaqian Mao, Wei Lin, Jixing Liang, Huibin Huang, Liantao Li, Junping Wen, Gang Chen

## Abstract

**Objective:** Our study aimed to investigate whether the metabolic indicators associated with non-communicable diseases (NCDs) in the general population have changed during the COVID-19 outbreak.

**METHODS:** This retrospective self-controlled study enrolled adult participants with metabolic indicators relate to NCDs followed at Fujian Provincial Hospital and Fujian Provincial Hospital South Branch. The metabolic indicators followed during January 1, 2020 and April 30, 2020, the peak period of the COVID-19 epidemic in China, were compared with the baseline value in the same period last year. Pared-samples T-test and Wilcoxon signed-rank test were performed to analyze the differences between paired data.

**Results:** The follow-up total cholesterol was significantly increased than that of the baseline (4.73 (4.05, 5.46) mmol/L vs 4.71 (4.05, 5.43) mmol/L, p=0.019; n=3379). Similar results were observed in triglyceride (1.29 (0.91, 1.88) vs 1.25 (0.87, 1.81) mmol/L, p<0.001; n=3381), uric acid (330.0 (272.0, 397.0) vs 327.0 (271.0, 389.0) umol/L, p<0.001; n=3364), and glycosylated hemoglobin (6.50 (6.10, 7.30) vs 6.50 (6.10, 7.20) %, p=0.013; n=532). No significant difference was observed in low density lipoprotein, body mass index and blood pressure.

**Conclusions:** Metabolic indicators associated with NCDs deteriorated in the COVID-19 outbreak. We should take action to prevent and control NCDs without delay.

## Introduction

At the end of 2019, the emergence of a new coronavirus named severe acute respiratory syndrome coronavirus 2 (SARS-CoV-2) caused the 2019 coronavirus disease (COVID-19) epidemic worldwide [1]. As of June 24th, there are over nine million confirmed cases of COVID-19 and almost 478,000 deaths globally [2]. Due to lack of effective vaccines and specific drugs, many countries have quickly adopted quarantine, social distance, community containment, and other measures to deal with this global public health emergency [3-5]. The Chinese government has implemented lockdown and social distance measures since Jan, 2020, and achieved preliminary success at the end of April [5, 6]. At present, the prevention and control of the COVID-19 epidemic in China has changed from emergency to normal, with lockdown status ended and social distance continued [7]. What we should pay attention to now is whether the risk and control of non-communicable diseases (NCDs) changed during the epidemic.

There is increasing clinical evidence that NCDs related to metabolic syndrome such as diabetes and hypertension were the most common comorbidities among COVID-19 patients, which can significantly increase the severity and mortality of SARS-CoV-2 infection [8-17]. In patients with COVID-19 and NCDs, blood pressure (BP) and blood glucose levels were associated with the prognosis [18, 19]. Recent clinical evidence suggested that obesity, the common risk factor for NCDs, was also independently associated with the progression and prognosis of patients with COVID-19 [20-23]. It was estimated that 45.4% of adults in the United States are at increased risk for complications from COVID-19 because of NCDs and its risk factors [24]. As the progression of COVID-19 is mainly dependent on the initial health status of an individual, the prevention and control of NCDs are of vital importance to reduce the harm of the COVID-19 epidemic in the future.

It’s worth noting that, lifestyles may change a lot in the face of the threat of COVID-19, thereby affecting the risk and control of NCDs [7, 25-27]. A recent analysis from mathematical modeling in India forecasted that lockdown will lead to a substantial increase in HbA1c and future diabetes-related complications [28]. People tend to eat and snack more unhealthy food under the condition of lockdown [7, 25, 27]. Meanwhile, time spent in sports activities decreased and screen time increased in the COVID-19 storm [26, 27]. These lifesty changes may lead to metabolic disorders related to NCDs. Furthermore, social distance and COVID-19 relevant negative news may adversely affect emotions, leading to behavior changes and physical health issues associated with NCDs [29]. Last but not least, routine management of NCDs has been scaled down during the peak period of the COVID-19 epidemic [30]. People at high risk for NCDs may have to delay their screening test, and patients with NCDs may discontinue treatment or change the treatment without the guidance of doctors, resulting in poor control.

To our knowledge, nevertheless, only a few studies focused on the risk of NCDs and its control during the COVID-19 outbreak. One study paid attention to weight change in Poland adults during the COVID-19 crises and found that almost 30% experienced weight gain [25]. The other studies focused on glycemic control in diabetes during the restrictions due to COVID-19 pandemics and unexpectly found no deterioration results [4, 31-33]. However, the sample size of those studies is small, and those who represent patients with nonautoimmune forms of diabetes are underrepresented.

NCDs are the leading causes of death worldwide, contributing to 73·4% of total deaths in 2017 [34]. The current double epidemic of NCDs and COVID-19 and their vicious circle relationship causes adverse health effects at the population level and at the same time places a wide burden on the health care system. The present retrospective self-controlled study aimed to analyze more metabolic indicators associated with NCDs in the general population on a large scale, offering the basis for the corresponding intervention strategy for the prevention and control of NCDs during the COVID-19 outbreak and the subsequent period.

## Results

### Demographic characteristics

The demographic characteristics of the studied population were presented in Table 1. More than three thousand participants were analyzed for total cholesterol (TC) (n=3379), low density lipoprotein (LDL) (n=3341), triglyceride (TG) (n=3381), and uric acid (UA) (n=3364). Middle-aged account for almost half. A total of 532 participants were analyzed for glycosylated hemoglobin (HbA1c) with most were middle-aged/aged and male. For body mass index (BMI)(n=445) and BP(n=463), more than four hundred of participants were analyzed, most of whom were young and female.

### Metabolic indicators

The comparison of baseline and follow-up metabolic indicators were shown in Table 2. The follow-up TC (4.73 (4.05, 5.46) mmol/L) was significantly increased than that of the baseline (4.71 (4.05, 5.43) mmol/L, p=0.019). Similar results were observed for TG (1.29 (0.91, 1.88) vs 1.25 (0.87, 1.81) mmol/L, p<0.001), UA (330.0 (272.0, 397.0) vs 327.0 (271.0, 389.0) umol/L, p<0.001) and HbA1c (6.50 (6.10, 7.30) vs 6.50 (6.10, 7.20) %, p=0.013). There was no significantly statistical difference between the follow up and baseline LDL (3.00 (2.40, 3.67) vs 3.04 (2.44, 3.70), p=0.169), BMI (22.86(20.72,24.88) vs 22.88(20.89,24.97), p=0.976), SBP (120.0 (111.0, 129.0) vs 120.0 (111.0, 130.0), p=0.896), and DBP (73.0 (67.0, 81.0) vs 73.0 (67.0, 81.0), p=0.724).

## Discussion

The main findings of the present study were that metabolic indicators including TC, TG, UA, and HbA1c were significantly increased during the outbreak of COVID-19. Nevertheless, no statistical difference was observed in LDL, BMI, and BP.

Our results provided evidence that metabolic indicators associated with NCDs deteriorated during the COVID-19 outbreak in China. With the initiation of the public health response to prevent the spread of COVID-19 since Jan, 2020 in China [5], the impacts of social distance and community containment during the epidemic on lifestyles may be one of the most important factors in the increase of TC, TG, UA, and HbA1c. People tend to eat and snack more under the condition of home quarantine [25]. As lockdown caused problems in the food supply chain and economic recession, the consumption of processed foods characterized by high fat, high sugar, and high salt increased due to easy availability, storage, and use [7, 25, 27, 35]. These dietary changes will lead to metabolic disorders related to NCDs in the COVID-19 outbreak and subsequent period. Meanwhile, the closure of gyms, swimming pools, and exercise clubs, in addition to home quarantine, will inevitably reduce opportunities to exercise [36]. A longitudinal study demonstrated that time spent in sports activities decreased and screen time increased in the COVID-19 storm[27]. This phenomenon is detrimental to health because physical activity is critical to control the symptoms and risk factors of NCDs [37]. Furthermore, fear of catching the virus, worrying about family, social isolation, financial pressure, rumors everywhere, and information overload lead to increased stress and anxiety levels, which will result in behavior changes and physical health issues associated with NCDs [29]. Last but not least, as precious healthcare resources were diverted towards the prevention and control of COVID-19 epidemics, and to avoid cross-contamination in hospitals, routine management of NCDs has been scaled down during the peak period of the COVID-19 epidemic in China [30]. Patients with NCDs who take angiotensin receptor blockers or angiotensin converting enzyme Inhibitors regularly may stop the treatment without the doctor’s guidance for fear that these medications will increase the risk of COVID-19 infection through up-regulation ACE2, and then lead to uncontrolled illness.

This is, as far as we know, the first study to explored blood lipid and uric acid levels among the general population during the outbreak of a public health emergency of international concern declared by the World Health Organization and proved increased blood lipid and uric acid. Though lack of clinical evidence about the association between hyperlipidemia/hyperuricemia and COVID-19, it’s seemed that metabolic disorders may indirectly affect the progression of COVID-19 through increase the risk of NCDs [38, 39]. In addition, lipids and cholesterol-rich membrane microdomains are essential for coronavirus entries in human cells and a high amount of intracellular cholesterol and fatty acids are essential for the formation of the replication complex of COVID-19 [40]. In this context, strengthen the screening and control of hyperlipidemia may have a double beneficial effect by reducing the NCD risk and interfering with COVID-19.

Our result that HbA1c was elevated in the COVID-19 storm provided further evidence for the prediction of previous arithmetic models [28]. However, previous studies focused on the glycemic control in diabetes during the restrictions due to COVID-19 pandemics found no deteriorated results [4, 31-33]. These different results may be explained as follows. Firstly, the sample size of previous studies is small (n=13, 33, 55, and 147), and patients with type 1 diabetes accounts for more than 90 percent of the studied population [4, 31-33]. Those representing patients with nonautoimmune forms of diabetes who are underrepresented in the previous study. Secondly, the patients in previous studies used both continuous glucose monitoring and platforms for remote data sharing, and were younger (median/average age <45 years old) than those in our study (median age 64 years old) [4, 31-33]. We speculate that the patients in previous studies may have better health literacy and compliance to avoid metabolic deteriorate in the COVID-19 outbreak. Our study enrolled the general population, including healthy people and patients with NCDs, with a larger sample size, and thus the conclusion may be more general. Last but not least, our research time is longer than that of previous studies. In the present study, it was a comparison between the peak period of COVID-19 epidemic and the same period of last year, not the short period of0020lockdown and pre-lockdown. The impact of the COVID-19 epidemic on NCDs and its risks is not only existed during the lockdown period, but also during the extended period of subsequent economic downturn and social distanc. It will probably take time to fully evaluate the impact of the COVID-19 outbreak on the NCDs risks and control.

It’s worth noting that, we did not find significant differences in BMI and BP between the time of the COVID-19 outbreak and the same period last year. These results may due to the small sample of BMI and BP of the participants in our study, and most of them are young people, who may have better health literacy and compliance to prevent metabolic deterioration in the COVID-19 storm.

The COVID-19 pandemic is expected to be long drawn. Meeting the challenge that metabolic indicators associated with NCDs deteriorated during the COVID-19 outbreak, we should take action to prevent and control NCDs without delay. We can focus on the following aspects to avoid the negative effects of the outbreak on general population. First of all, as the primary care system plays an important role in the prevention and control of NCDs, a hierarchical medical system should be further implemented and medical education for general practitioners should be enhanced. General practitioners should not only focus on the prevention and control of infectious disease but also NCDs with the context of their community. Metabolic indicators associated with NCDs should be further monitored by general practitioners during the outbreak in order to return them to normal as soon as possible, and thus reduce the morbidity and mortality rates of NCDs. Telemedicine consults should be implemented where possible and home delivery services should be arranged for essential medications. Secondly, with the development of 5th generation wireless systems, high-quality massive open online courses on health education should be developed by health workers and thus promote healthy behaviors at home. Health education and health-promoting behaviors have been proven beneficial for metabolic management [41, 42]. In addition, the government should take measures to ensure easy access to healthy foods for the population and discourage the use or consumption of unhealthy products. Last but not least, information sharing should be enhanced and proper psychological guidance should be implemented to help alleviate the psychological impact of the pandemic, and thus reduce physical health issues associated with NCDs [29].

The main limitation of our study is that it only reflected changes in metabolic indicators associated with NCDs during the COVID-19 outbreak. With no detailed information relating to particular changes in eating habits, physical activity, and psychological situation from the participants, we could not confirm which one lead to the deterioration of the metabolic indicators most. In addition, these data refered to an adult cohort including healthy people and patients with NCDs. Information of medical history that may affect the results were not recorded. More studies are necessary to fully evaluate the impact that COVID-19 has had on the health status at population level, and it is of vital importance to collect information from now on to better prevent and control NCDs.

In conclusion, metabolic indicators associated with NCDs deteriorated in the COVID-19 outbreak. It is a crucial time to strengthen action on prevention and control of NCDs to minimize the morbidity and mortality rates of COVID-19 in the short-term and reduce total morbidity and mortality rates of NCDs in the long-term, avoiding adding on to the burden of countries’ healthcare systems.

## Methods

This retrospective study was approved by the Ethics Committee of Fujian Provincial Hospital and Fujian Provincial Hospital South Branch. Due to the retrospective nature of the study, informed consent was waived. Participants’ information has been anonymized since the data collection stage.

Individuals aged 18 years old and above with metabolic indicators associated with NCDs followed at Fujian Provincial Hospital and Fujian Provincial Hospital South Branch were enrolled in our study. Emergency patients and inpatients were excluded. Metabolic data including TC, LDL, TG, UA, HbA1c, BMI, SBP and DBP followed during January 1, 2020 and April 30, 2020, the peak period of COVID-19 epidemic in China[6], were compared with the baseline value in the same period in 2019. Participants aged 18-44, 45-64, and >65 years old were regarded as young, middle-aged, and aged.

All statistical analyses were conducted with SPSS Statistics for Windows, Version 25.0 (Armonk, NY: IBM Corp). Data are presented as the number (percentage) for categorical variables and the median (interquartile range (IQR)) for continuous variables that are not normally distributed. Pared-samples T-test and Wilcoxon signed-rank test were performed to check the differences of paired data that are normally distributed and abnormally distributed, respectively. All tests were two-sided and p<0.05 was considered statistically significant.

## Data Availability

All data included in this study are available upon request by contact with the corresponding author.

## Abbreviations

ACE2: angiotensin-converting-enzyme 2
BMI: body mass index
BP: blood pressure
COVID-19: 2019 coronavirus disease
DBP: diastolic blood pressure
HbA1c: glycosylated hemoglobin
IQR: interquartile range
LDL: low density lipoprotein
NCDs: non-communicable diseases
SARS-CoV-2: severe acute respiratory syndrome coronavirus 2
SBP: systolic blood pressure
TC: total cholesterol
TG: triglyceride
UA: uric acid

## Author Contributions

Ting Xue wrote the first draft of the manuscript. Ting Xue, Lizhen Xu, and Yaqian Mao collected the data and conducted analyses. Gang Chen revised the manuscript, reviewed the results, and contributed to the discussion. Gang Chen conceived of the research idea. Gang Chen is the guarantors of this work and, as such, had full access to all the data in the study and takes responsibility for the integrity of the data and the accuracy of the data analysis.

## Acknowledgments

We greatly appreciate the efforts of all the hospital employees and their families at the Fujian Provincial Hospital and Fujian Provinvial Hospital Soutu Branch, who are working tirelessly during this outbreak.

## References

1. Zabetakis I, Lordan R, Norton C, Tsoupras A. COVID-19: The Inflammation Link and the Role of Nutrition in Potential Mitigation. Nutrients. 2020; 12.

2. John Hopkins University. John Hopkins University & Medicine: Coronavirus Resource Center. Available online: https://coronavirus.jhu.edu/map.html (accessed on 24 Jun 2020).

3. Hall G, Laddu DR, Phillips SA, Lavie CJ, Arena R. A tale of two pandemics: How will COVID-19 and global trends in physical inactivity and sedentary behavior affect one another? Prog Cardiovasc Dis. 2020.

4. Tornese G, Ceconi V, Monasta L, Carletti C, Faleschini E, Barbi E. Glycemic Control in Type 1 Diabetes Mellitus During COVID-19 Quarantine and the Role of In-Home Physical Activity. Diabetes Technol Ther. 2020; 22: 462–7.

5. Wilder-Smith A, Freedman DO. Isolation, quarantine, social distancing and community containment: pivotal role for old-style public health measures in the novel coronavirus (2019-nCoV) outbreak. J Travel Med. 2020.

6. National Health Commission of the People’s Republic of China, Available online : http://www.nhc.gov.cn/xcs/yqtb/list_gzbd.shtml (accessed on 24 Jun 2020)..

7. Bureau of Disease Control and Prevention of the People’s Republic of China, Available online : http://www.nhc.gov.cn/jkj/s5899tg/202005/aa359e8f8e2648e7a474e7b38aacb4b0.shtml (accessed on 24 Jun 2020).

8. Wang B, Li R, Lu Z, Huang Y. Does comorbidity increase the risk of patients with COVID-19: evidence from meta-analysis. Aging (Albany NY). 2020; 12: 6049–57.

9. Yang J, Zheng Y, Gou X, Pu K, Chen Z, Guo Q, Ji R, Wang H, Wang Y, Zhou Y. Prevalence of comorbidities and its effects in patients infected with SARS-CoV-2: a systematic review and meta-analysis. Int J Infect Dis. 2020; 94: 91–5.

10. Zhang J, Wu J, Sun X, Xue H, Shao J, Cai W, Jing Y, Yue M, Dong C. Association of hypertension with the severity and fatality of SARS-CoV-2 infection: A meta-analysis. Epidemiol Infect. 2020; 148: e106.

11. Zheng Z, Peng F, Xu B, Zhao J, Liu H, Peng J, Li Q, Jiang C, Zhou Y, Liu S, Ye C, Zhang P, Xing Y, et al. Risk factors of critical & mortal COVID-19 cases: A systematic literature review and meta-analysis. J Infect. 2020.

12. Parohan M, Yaghoubi S, Seraji A, Javanbakht MH, Sarraf P, Djalali M. Risk factors for mortality in patients with Coronavirus disease 2019 (COVID-19) infection: a systematic review and meta-analysis of observational studies. Aging Male. 2020: 1–9.

13. Liu H, Chen S, Liu M, Nie H, Lu H. Comorbid Chronic Diseases are Strongly Correlated with Disease Severity among COVID-19 Patients: A Systematic Review and Meta-Analysis. Aging Dis. 2020; 11: 668–78.

14. Li X, Guan B, Su T, Liu W, Chen M, Bin Waleed K, Guan X, Gary T, Zhu Z. Impact of cardiovascular disease and cardiac injury on in-hospital mortality in patients with COVID-19: a systematic review and meta-analysis. Heart. 2020.

15. Li B, Yang J, Zhao F, Zhi L, Wang X, Liu L, Bi Z, Zhao Y. Prevalence and impact of cardiovascular metabolic diseases on COVID-19 in China. Clin Res Cardiol. 2020; 109: 531-8.

16. Jain V, Yuan JM. Predictive symptoms and comorbidities for severe COVID-19 and intensive care unit admission: a systematic review and meta-analysis. Int J Public Health. 2020: 1–14.

17. Tadic M, Cuspidi C, Sala C. COVID-19 and diabetes: Is there enough evidence? J Clin Hypertens (Greenwich). 2020.

18. Wang Z, Du Z, Zhu F. Glycosylated hemoglobin is associated with systemic inflammation, hypercoagulability, and prognosis of COVID-19 patients. Diabetes Res Clin Pract. 2020; 164: 108214.

19. Vicenzi M, Di Cosola R, Ruscica M, Ratti A, Rota I, Rota F, Bollati V, Aliberti S, Blasi F. The liaison between respiratory failure and high blood pressure: evidence from COVID-19 patients. Eur Respir J. 2020.

20. Klang E, Kassim G, Soffer S, Freeman R, Levin MA, Reich DL. Morbid Obesity as an Independent Risk Factor for COVID-19 Mortality in Hospitalized Patients Younger than 50. Obesity (Silver Spring). 2020.

21. Kalligeros M, Shehadeh F, Mylona EK, Benitez G, Beckwith CG, Chan PA, Mylonakis E. Association of Obesity with Disease Severity among Patients with COVID-19. Obesity (Silver Spring). 2020.

22. Hajifathalian K, Kumar S, Newberry C, Shah S, Fortune B, Krisko T, Ortiz-Pujols S, Zhou XK, Dannenberg AJ, Kumar R, Sharaiha RZ. Obesity is associated with worse outcomes in COVID-19: Analysis of Early Data From New York City. Obesity (Silver Spring). 2020.

23. Gao F, Zheng KI, Wang XB, Sun QF, Pan KH, Wang TY, Chen YP, Targher G, Byrne CD, George J, Zheng MH. Obesity Is a Risk Factor for Greater COVID-19 Severity. Diabetes Care. 2020.

24. Adams ML, Katz DL, Grandpre J. Population-Based Estimates of Chronic Conditions Affecting Risk for Complications from Coronavirus Disease, United States. Emerg Infect Dis. 2020; 26.

25. Sidor A, Rzymski P. Dietary Choices and Habits during COVID-19 Lockdown: Experience from Poland. Nutrients. 2020; 12.

26. Martinez-Ferran M, de la Guía-Galipienso F, Sanchis-Gomar F, Pareja-Galeano H. Metabolic Impacts of Confinement during the COVID-19 Pandemic Due to Modified Diet and Physical Activity Habits. Nutrients. 2020; 12.

27. Pietrobelli A, Pecoraro L, Ferruzzi A, Heo M, Faith M, Zoller T, Antoniazzi F, Piacentini G, Fearnbach SN, Heymsfield SB. Effects of COVID-19 Lockdown on Lifestyle Behaviors in Children with Obesity Living in Verona, Italy: A Longitudinal Study. Obesity (Silver Spring). 2020.

28. Ghosal S, Sinha B, Majumder M, Misra A. Estimation of effects of nationwide lockdown for containing coronavirus infection on worsening of glycosylated haemoglobin and increase in diabetes-related complications: A simulation model using multivariate regression analysis. Diabetes Metab Syndr. 2020; 14: 319–23.

29. Abbas AM, Fathy SK, Fawzy AT, Salem AS, Shawky MS. The mutual effects of COVID-19 and obesity. Obes Med. 2020; 19: 100250.

30. National Health Commission of the People’s Republic of China, Available online : https://www.cma.org.cn/art/2020/3/14/art_2928_33636.html (xaccessed on 24 Jun 2020).

31. Beato-Víbora PI. No deleterious effect of lockdown due to COVID-19 pandemic on glycaemic control, measured by glucose monitoring, in adults with type 1 diabetes. Diabetes Technol Ther. 2020.

32. Bonora BM, Boscari F, Avogaro A, Bruttomesso D, Fadini GP. Glycaemic Control Among People with Type 1 Diabetes During Lockdown for the SARS-CoV-2 Outbreak in Italy. Diabetes Ther. 2020: 1–11.

33. Maddaloni E, Coraggio L, Pieralice S, Carlone A, Pozzilli P, Buzzetti R. Effects of COVID-19 Lockdown on Glucose Control: Continuous Glucose Monitoring Data From People With Diabetes on Intensive Insulin Therapy. Diabetes Care. 2020.

34. Global, regional, and national age-sex-specific mortality for 282 causes of death in 195 countries and territories, 1980-2017: a systematic analysis for the Global Burden of Disease Study 2017. Lancet. 2018; 392: 1736–88.

35. Oliveira TC, Abranches MV, Lana RM. Food (in)security in Brazil in the context of the SARS-CoV-2 pandemic. Cad Saude Publica. 2020; 36: e00055220.

36. Palmer K, Monaco A, Kivipelto M, Onder G, Maggi S, Michel JP, Prieto R, Sykara G, Donde S. The potential long-term impact of the COVID-19 outbreak on patients with non-communicable diseases in Europe: consequences for healthy ageing. Aging Clin Exp Res. 2020: 1–6.

37. Dekker J, Buurman BM, van der Leeden M. Exercise in people with comorbidity or multimorbidity. Health Psychol. 2019; 38: 822–30.

38. Mortada I. Hyperuricemia, Type 2 Diabetes Mellitus, and Hypertension: an Emerging Association. Curr Hypertens Rep. 2017; 19: 69.

39. Bertolotti M, Maurantonio M, Gabbi C, Anzivino C, Carulli N. Review article: hyperlipidaemia and cardiovascular risk. Aliment Pharmacol Ther. 2005; 22 Suppl 2: 28–30.

40. Scicali R, Di Pino A, Piro S, Rabuazzo AM, Purrello F. May statins and PCSK9 inhibitors be protective from COVID-19 in familial hypercholesterolemia subjects? Nutr Metab Cardiovasc Dis. 2020.

41. Dempsey PC LR, Sethi P, Sacre JW, Straznicky NE, Cohen ND, Cerin E, Lambert GW, Owen N, Kingwell BA, Dunstan DW. Benefits for Type 2 Diabetes of Interrupting Prolonged Sitting With Brief Bouts of Light Walking or Simple Resistance Activities. Diabetes Care 2016; 39: 964–72.

42. Fan R, Xu M, Wang J, Zhang Z, Chen Q, Li Y, Gu J, Cai X, Guo Q, Bao L, Li Y. Sustaining Effect of Intensive Nutritional Intervention Combined with Health Education on Dietary Behavior and Plasma Glucose in Type 2 Diabetes Mellitus Patients. Nutrients. 2016; 8.

